# Rapid and prolonged antidepressant and antianxiety effects of psilocybin, lysergic acid diethylamide, ayahuasca, and 3, 4-methylenedioxy-methamphetamine. A systematic review and meta-analysis of randomized controlled trials

**DOI:** 10.1101/2024.06.17.24308787

**Authors:** Dimy Fluyau, Vasanth Kattalai Kailasam, Neelambika Revadigar

**Affiliations:** Emory University School of Medicine, Atlanta, GA, USA; Mercyhealth Pain Center–Crystal Lake, Mercyhealth Hospital and Physician Clinic–Crystal Lake, Mercyhealth Pain Center–Harvard L 875 S. Route 31, Crystal Lake, IL 60014, USA; Santa Clara Valley Medical Center, 751 S Bascom Ave, San Jose, CA 95128, USA

**Keywords:** Hallucinogens, major depressive disorder, anxiety disorder, systematic review, meta-analysis

## Abstract

**Background:** Hallucinogens attract research as alternatives to the commonly used medications to treat major depressive and anxiety disorders.

**Aims:** Assess hallucinogens’ efficacy for managing depressive and anxiety symptoms and evaluate their safety profiles.

**Method:** In five databases, we searched for randomized controlled trials of hallucinogens targeting depressive and anxiety symptoms. We performed a meta-analysis using a random effects model when data permitted it. The protocol of the review is registered in PROSPERO; CRD42022341325.

**Results:** Psilocybin produced a rapid and sustained reduction in depressive and anxiety symptoms in patients with major depressive disorder, severe, and in patients with life-threatening cancer. A decrease in depressive symptoms was observed with 3, 4-methylenedioxymethamphetamine (MDMA), primarily in patients with life-threatening cancer, autism spectrum disorder, and post-traumatic stress disorder. MDMA reduced social anxiety symptoms. However, MDMA’s effect size was either negligible or negative for anxiety symptoms overall. Ayahuasca reduced depressive symptoms in individuals with treatment-resistant major depressive and personality disorders. Lysergic acid diethylamide (LSD) reduced anxiety symptoms in individuals with life-threatening cancer.

Psilocybin’s adverse effects were noticeable for elevated blood pressure, headaches, and panic attacks. For MDMA, elevated blood pressure, headaches, panic attacks, and feeling cold were noticeable.

**Conclusions:** Psilocybin, MDMA, ayahuasca, and LSD appear to have the potential to reduce depressive and anxiety symptoms. Adverse effects are noticed. Rigorous randomized controlled studies with larger sample sizes utilizing outcome measures instruments with better reliability and validity are warranted.

## Introduction

Major depressive and anxiety disorders are psychiatric disorders that can cause severe impairment in functioning. Pharmacotherapy based on reuptake inhibitors (serotonin and norepinephrine), receptor antagonists (serotonin 5-HT2 receptors and dopamine D2 receptor), enzyme inhibitors (monoamine oxidase inhibitors A and B) or reversible enzyme inhibitors (monoamine oxidase inhibitors A), enzyme releasers (dopamine transporter, norepinephrine) as monotherapy or in combination with other pharmacological treatments to treat the disorders take time to produce a beneficial effect. Many patients remain ill or develop treatment resistance. Technical developments on faulty brain circuits and neuroplasticity have led to the advancement of new strategies for identifying novel agents with the potential to improve treatment. Hallucinogens are one of the apparent targets, likely because of their possible immediate and long-lasting efficacy. Esketamine, the enantiomer of ketamine, is accessible for treatment-resistant major depressive disorder in the United States of America (USA). Its use is restricted due to the potential for abuse and misuse. Around 51% of 6381 people who reported past-year hallucinogen use in the 2016-2018 National Survey on Drug Use and Health abused lysergic acid diethylamide (LSD), psilocybin (mushroom), or 4-methylenedioxymethamphetamine (MDMA) (1). Documentation of hallucinogen risks for addiction is scant, and their adverse profile pictures mostly the term: no serious adverse effects.

Psychedelics (LSD, mescaline, ayahuasca, psilocybin), dissociatives (phencyclidine)(PCP), dextromethorphan (DMX), ketamine), deliriants (atropine, scopolamine), and kappa-opioid agonists (Salvinorin A) and entactogens are notable hallucinogens (2,3) that alter behavior, mood, thought, and perception (2). Psilocybin, ayahuasca, and LSD have been the subjects of study for the management of depressive and anxiety symptoms (3, 4), as has the entactogen MDMA for post-traumatic stress disorder (PTSD) and life-threatening illness-related anxiety (6, 7).

Psilocybin activates 5HT2A-R and 5HT1A-R (8, 9) within default-mode network DMN-associated brain regions, inhibits the activity of adenylate cyclase, decreases protein kinase A-mediated extracellular Ca2+ influx (10) and stimulates glutamate in contrast to γ-aminobutyric acid (GABA) release (11). Psilocybin, consumed as mushrooms, is sometimes sold on the street as phencyclidine (PCP). 10 mg (about the weight of a grain of table salt) of psilocybin per gram of mushroom is said to cause agitation, hallucinations, dizziness, weakness, and anxiety (12).

The median lethal dose (LD50) for psilocybin in rats and mice is 280–285 mg (about the weight of ten grains of rice)/kg, while in rabbits, it is 12.5 mg (about half the weight of a grain of rice)/kg (13). It is presumed that psilocybin use and harm are low compared to illicit substances (14).

LSD interacts directly with 5-HT1A, 5-HT2A, 5-HT2C, D2, and α2 receptors and indirectly with glutamate through N-methyl-d-aspartate (NMDA) receptors (15). LSD (also known as acid on the street) can cause panic attacks, horrific hallucinations, horrific illusions), long-term psychosis, and post-hallucinogen perception disorder (16, 17). LSD LD50 values range from 0.3 mg/kg intravenously (i.v.) in rabbits to 16.5 mg/kg i.v. in rats and 46-60 mg/kg i.v. in mice (18). Between 2002 and 2018, the National Survey on Drug Use and Health found a rise in LSD use among those with a bachelor’s degree, single status, antisocial behavior, and co-occurring mental health and drug abuse disorders (19).

Ayahuasca interacts with dopamine, serotonin (5HT2A, 5HT1A, and 5HT2C), and norepinephrine through its monoamine oxidase inhibitors activity (20, 21, 22, 23), respectively. Ayahuasca has hormonal effects, specifically elevating prolactin levels, cortisol, and growth hormone (24). There have been reports of toxicity in humans and animals (25). Wiltshire and colleagues reported the death of a man who consumed ayahuasca, psilocybin, cannabis, and papaver seeds (26). Lima and colleagues noted that administering ayahuasca to rats at a dose of 2.5 mg/kg caused a reduction in locomotion by 44% and a reduction in vertical exploration by 62% (27). According to Pitol and colleagues, providing Wistar rats with 4 mL/kg over 14 days led to arterial hypertrophy (28). Abuse of ayahuasca was evaluated using the Addiction Severity Index by Fábregas and colleagues, and the researchers concluded that ayahuasca does not have negative psychosocial repercussions consistent with other addictive drugs (29).

MDMA releases serotonin (30), dopamine, and norepinephrine (31), as well as oxytocin, vasopressin, and cortisol (32). The LD50 of MDMA in animals ranges from about 100 to 300 mg/kg. Most fatalities involving MDMA are attributable to dehydration and concurrent drug intoxication (33, 34, 35). MDMA is frequently abused for recreational purposes (36). It is reported that MDMA has a less intense reinforcing effect than other substances used for recreational purposes but still carries the risk of addiction (37).

Findings from open-label and randomized controlled trials (RCTs) signaled that hallucinogens can manage depressive and anxiety symptoms and are well-tolerated. Literature reviews on hallucinogens’ benefits focused on psilocybin, and most reviews clustered different hallucinogens in one group. Also, literature reviews showed less or no emphasis on hallucinogens’ possible long-term benefits. Li and colleagues focused on psilocybin for depressive symptoms (38). Romeo and colleagues evaluated the combined effects of psychedelics (psilocybin, ayahuasca, and LSD) on depressive symptoms (39). Leger and colleagues assessed the methodological differences in outcomes in using psychedelics for anxiety and depressive disorders (40). Goldberg and colleagues focused on the antianxiety and antidepressant effects of psilocybin before administration and before follow-up (41). Vargas and colleagues examined psilocybin only for depressive and anxiety symptoms (42). Galvo-Coelho and colleagues investigated the acute, medium-term, and long-term antidepressant effects of psilocybin and ayahuasca (43).

Pharmacological differences between individual hallucinogens can be substantive, as can differences in their clinical responses. Thus, we individually assessed hallucinogens’ rapid (two weeks) and sustained (six months and up) responses to reduce depressive and anxiety symptoms and their safety profiles. To mitigate discrepancies between outcome measures, instruments that quantified hallucinogens’ clinical responses were used in clinician-reported or self-reported outcome measures.

## Method

### Criteria for considering studies

#### Types of studies

We included relevant RCTs in our investigation regardless of the publication date, country of origin, language, or outcomes. We excluded open-label trials.

#### Types of participants

Adults 18 –years–old or older with or without any psychiatric history with or without a medical illness.

### Types of interventions

#### Experimental intervention

Any hallucinogen that alleviates depressive or anxiety symptoms. Control intervention

The control intervention can be a placebo or any active pharmacological agent as a comparator.

### Types of outcome measures

#### Clinical response

Reduction in clinician-rated depressive and anxiety symptoms or self-reported reduction.

#### Adverse events

Any medical or neuropsychiatric incident related to hallucinogens.

### Types of settings

The settings were not limited and included medical centers, outpatient clinics, academic universities, or hospitals.

### Search methods for identification of studies

#### Electronic searches

The review was based on the Preferred Reporting Items for Systematic Reviews and Meta-Analyses (PRISMA) criteria (44). It is listed on the International Prospective Register of Systematic Reviews (PROSPERO; CRD42022341325). Articles published from inception to June 2022 were searched in the following databases: Academic Search Complete (1976–2022), Scopus (1998–2022), Embase, PubMed (1975–2022), and Google Scholar (2018–2022). Search terms were for example: hallucinogens+and+depression+and+randomized+controlled+trials;psychedelics+and+depression+and+randomized+controlled+trials.

### Data collection and analysis

Two authors (DF and VK) independently screened the titles of all studies obtained by the search strategy, excluded all irrelevant articles, and then retained potentially relevant studies. NR solved disagreements between DF and VK.

The following data were extracted:

- Publication status, title, authors’ names, source, country, and year of publication.
- Trial characteristics: design and setting.
- Interventions: type of pharmacological and control intervention, dose, and duration.
- Number of participants, age, gender, loss to follow-up, and race.
- Outcomes.

### Evaluation of the methodological quality of randomized controlled trials, bias risk, and heterogeneity

Methodological and risk of bias considerations are based on the Cochrane Handbook for Systematic Reviews of Interventions guidelines (81). The JADAD scale analyzed the methodological quality of clinical trials independently, and each trial was assigned a JADAD score. The JADAD scale ranges from 0 to 5, with trials scoring 3 or above regarded to be of high quality (82). Using the I^2^ statistic, statistical heterogeneity between studies was evaluated (45).

I^2^ = 0% to 40%: might not be important.

I^2^ =30% to 60%: may represent moderate heterogeneity. I^2^ =50% to 90%: may represent substantial heterogeneity. I^2^ =75% to 100%: considerable heterogeneity.

### Data synthesis

A pooled effect size analysis was performed when the combined studies reached at least two. In such cases, data were synthesized by using random-effects frequentist meta-analysis. Results were quantified and interpreted by the standardized mean difference (Cohen’s d) and 95% confidence interval. A Cohen’s d =0.2 was interpreted as a small effect size, 0.5 as moderate, and 0.8 as large. 46 Results yielded a p-value of .05, and below were considered statistically significant. Meta-analyses were conducted in Jeffreys’s Amazing Statistics Program, JASP Team (2020). JASP (Version 0.14.1) [Computer software] (University of Amsterdam Nieuwe Achtergracht 129B Amsterdam, The Netherlands and Open-source, Cross-platform Software for Ecological and Evolutionary Meta-analysis (OpenMEE). We performed a qualitative summary. when a meta-analysis was not feasible; and provided the effect size result.

A meta-regression analysis was conducted when there were two or more studies on a particular hallucinogen utilizing moderators such as age, hallucinogen dosage, sample size, gender, dropout rate, adverse effects, and race. Lastly, a table summary of the selected variables was presented, comprising demographics, setting, study design, and outcomes.

No ethical approval or written informed consent was required for this review as no patient specific information was gathered or evaluated.

## Results

### Description of studies

After screening, the search strategy yielded 49 studies (Fig. 1). Sixteen of them qualified for inclusion in the investigation. We excluded three trials in which depressive or anxiety results were executed on the open-label design (47, 48, 49), and one trial conducted in a naturalistic setting (50). We included 12 trials (51, 52, 53, 54, 55, 56, 57, 58,59,60,61, 62), ten of which provided data, allowing us to conduct a meta-analysis. Table 1 summarizes the individual characteristics of the studies included. More than half of the twelve trials took place in the United States of America (USA). Half of them were cross-over trials. Psychotherapy was provided in nine of the twelve studies. The study locations varied, but there was a trend toward academic/university and outpatient settings. Trial participants were diagnosed with life-threatening cancer, moderate to severe major depressive disorder, treatment-resistant major depressive disorder, post-traumatic stress disorder (PTSD), and autism spectrum disorder. For all the studies, the mean(μ) sample size was 32.75 with a standard deviation(σ) of 22.24, a μ age of 38.75, σ 13.43, psilocybin dosages μ 25.25 mg, σ 3.19 per 70kg, MDMA dosages μ116 mg, σ 10.55, LSD dosage 200 μg, and ayahuasca 25 mg/70 kg. Five studies involved psilocybin, and among them, the study of Davis and colleagues (51) included a waiting-list period for which data were analyzed in the long-term and were based on Gukasyan and colleagues’ analysis (63).

**Fig 1.**
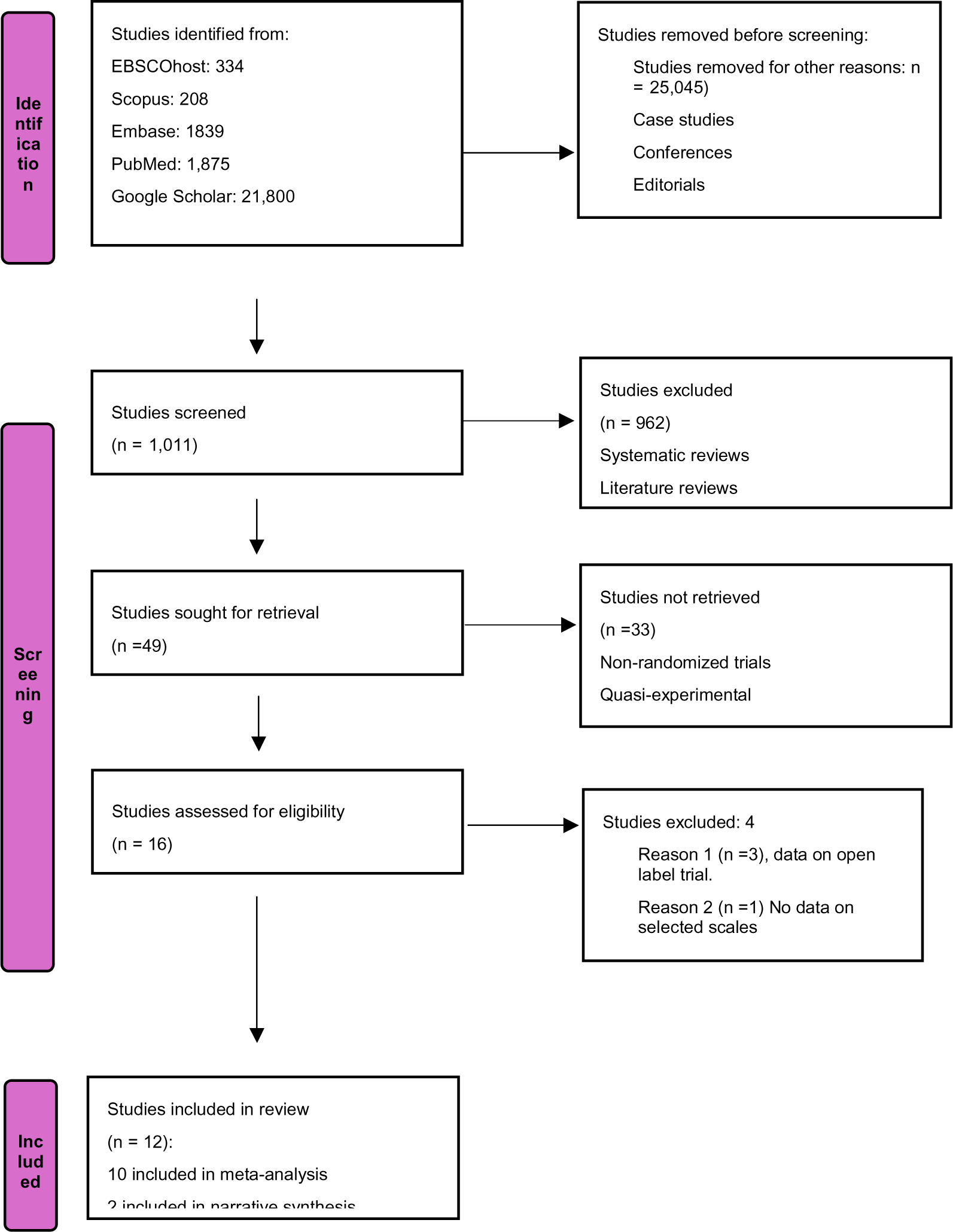
Flow diagram of the studies selection process.

**Table 1.**
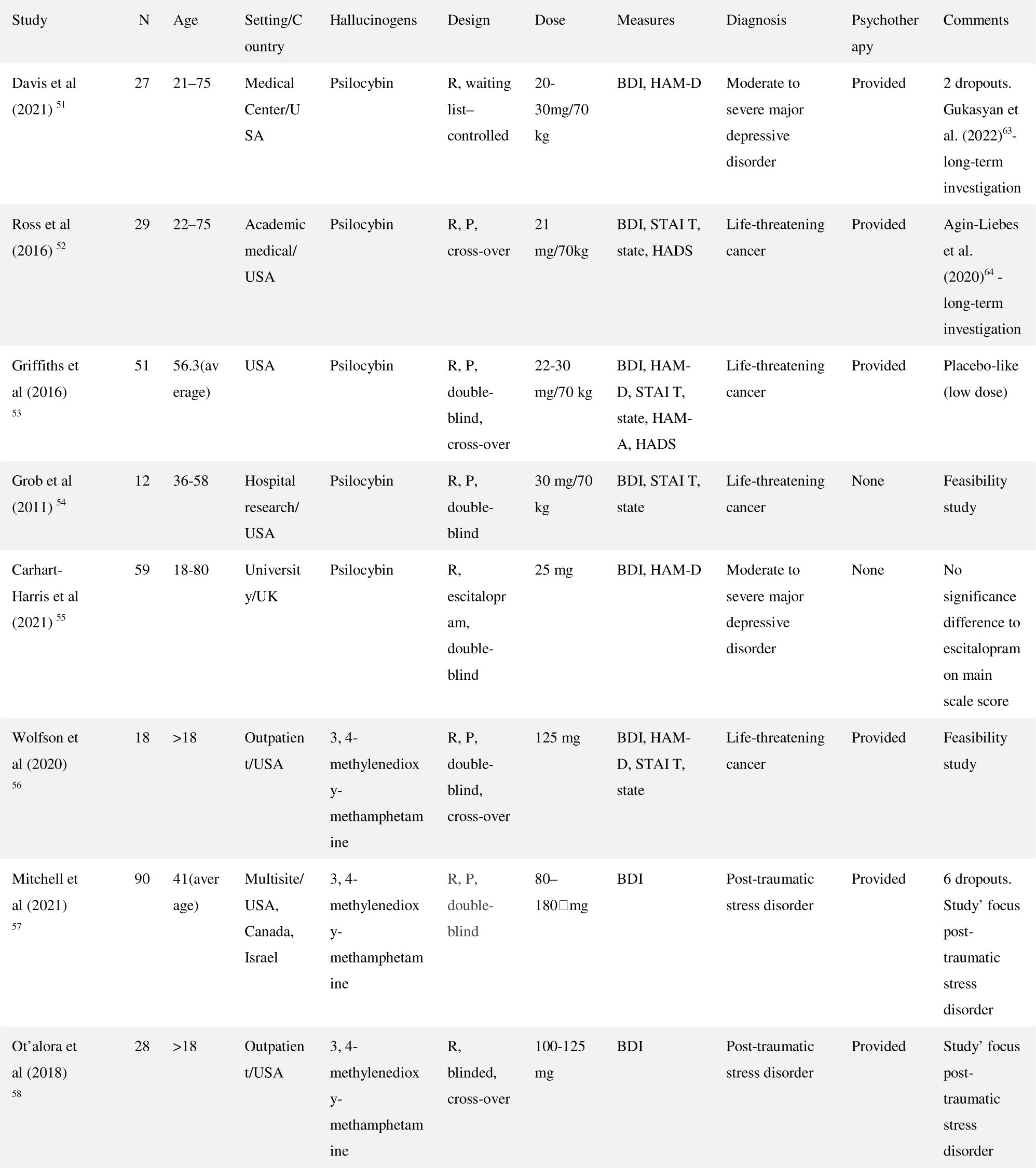
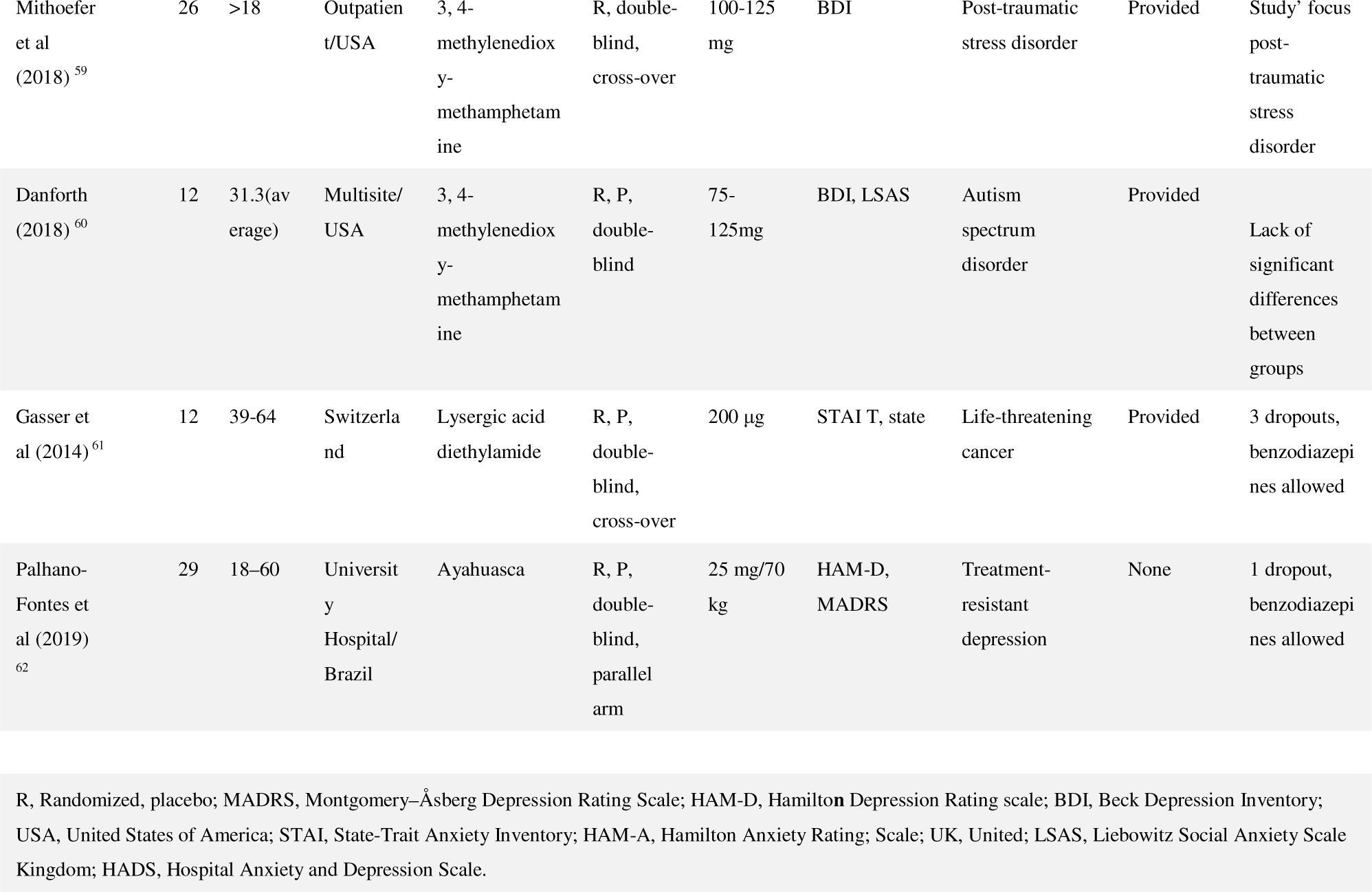
Characteristics of the included studies

Psilocybin’s long-term benefit from Ross and colleagues 52 was investigated through data from Agin-Liebes and colleagues (64).

### Risk of bias in included studies

The average JADAD score for all studies was 4.18. Overall, we judged that the twelve studies selected carried a minimal risk of bias.

### Synthesis of results

#### Depressive symptoms -Clinician-rated measures

Psilocybin. Pooled data at six months in patients with moderate and severe major depressive and life-threatening cancer showed that psilocybin (20-30mg/70 kg) significantly reduced Hamilton Depression Rating Scale (HAM-D) scores (d = 2.814, 95 % CI = 2.210 to 3.418, P= 0.001, I^2^ = 0%). A pooled analysis was not feasible. At week six, psilocybin (25 mg) outperformed escitalopram in reducing HAM-D scores in individuals with moderate to severe depression (not corrected for multiple comparisons) (d 2.3949; 95% CI= 1.7261 to 3.0636).

Ayahuasca. A pooled analysis was not feasible. When ayahuasca (0.36 mg/kg) was compared to a placebo, the HAM-D (d = 0.8495, 95% CI= 0.21 to 1.75) and the Montgomery-Asberg Scale for Depression (MADRS) scores (d = 1.49, 95% CI = 0.67 to 2.32) dropped rapidly at week one in individuals with treatment-resistant major depressive disorder with a high level of personality disorder.

#### Depressive symptoms - Self-rated measures

Psilocybin. In individuals with life-threatening cancer, pooled analysis indicated that psilocybin (20-30mg/70 kg) decreased the Beck Depression Inventory (BDI) score significantly in two weeks compared to placebo (d = 1.023, 95% CI = 0.424 to 1.622, P = 0.001, I^2^ = 0%). At week six, in individuals with moderate to severe depression, as a secondary outcome, psilocybin (25 mg) fared better than escitalopram in reducing the BDI score (d = 1.0431, 95% CI = 0.4941 to 1.5431) (not corrected for multiple comparisons). In moderate to severe depression with life-threatening cancer, pooled analysis indicated that the BDI score reduction was maintained at six months (d = 1.907, 95% CI= 1.482 to 2.333, P = 0.001, I^2^ = 0%). At six months in individuals with life-threatening cancer, a decrease in Hospital Anxiety and Depression Scale (HADS depression) was also seen (d = 2,311, 95% CI = 0.987 to 3.634, P = 0.001). However, the real effect may differ due to high heterogeneity (I^2^ = 78,474).

MDMA. Data extracted on MDMA originated from individuals with life-threatening cancer, autistic adults, and PTSD. Pooled analysis suggested that MDMA (75–180 mg) could lower BDI (d = 0.882, 95% CI = 0.467 to 1.296, P = 0.001, I^2^= 0%) from two months to twelve months compared to treatment at baseline.

#### Anxiety symptoms-Clinician-rated measures

Psilocybin. Pooled analysis in individuals with life-threatening cancer, who stayed in treatment for up to six months, indicated that psilocybin significantly reduced the Hamilton Anxiety Rating Scale (HAM-A) score (d = 2.853, 95% CI = 1.638 to 4.068, P < 0.001). The actual effect may differ due to high heterogeneity (I^2^ = 68.76%).

MDMA. A pooled analysis was not viable. In autistic adults, at 6-month follow-up, MDMA (75-125 mg) appeared to induce a decline in Liebowitz Social Anxiety Scale total scores (d = 1.4826, 95% CI = 0.1438 to 2.8213).

#### Anxiety symptoms-Self-rated measures

Psilocybin. In life-threatening cancer from baseline to week two, psilocybin (20-30mg/70 kg) compared to placebo significantly reduced State-Trait Anxiety Inventory (STAI) (d = 1.158, 95% CI = 0.538 to 1.778, P < 0.001, I^2^ = 0%) and STAI-State scores (d = 1.066, 95% CI = 0.33 to1.798, P < 0.004, I^2^ = 33.75%). At six months, in individuals with life-threatening cancer psilocybin (21-30 mg/70kg) significantly reduced both STAI-Trait (d =1.404, 95% CI = 0.818 to 1.990, P < 0.001, I^2^ = 0%) and STAI-State scores (d = 1.310, 95% CI= 0.654 to 1.965, P < 0.001, I^2^ = 0%). However, psilocybin’s effect size reduction on the Hospital Anxiety and Depression Scale (HADS anxiety) was small and non-statistically significant (d = 0.219, 95% CI = -0.400 to 0.838, P = 0.488) with moderate heterogeneity (I^2 =^ 47.666%).

MDMA. At 6 months, in autistic adults and in individuals with life-threatening cancer MDMA (75-125 mg)’s reduction in both STAI-Trait (d= 0.462, 95% CI = -0.737 to 1.662, P = 0.450,) and STAI-State scores (d = -0.139, 95% CI =-1.416 to 1.138, P = 0.831,) was not statistically significant and with substantial heterogeneity (I^2^ = 54.491% and I^2^ = 59.704%, respectively). LSD In individuals with life-threatening cancer who received LSD (200 μg) over two months compared to a placebo, there was an indication that LSD could reduce STAI-Trait (d= 1.1, 95%CI= 0.86 to 3.06) and STAI-State scores (d = 1.2, 95% CI 0.76 to 3.16).

### Meta-regression

Potential moderators of psilocybin and MDMA’s therapeutic effects including age, hallucinogen dosage, sample size, gender, dropout rate, adverse effects, and race had no meta effect on results.

### Adverse effects

Twenty-two types of adverse effects were surveyed. On average, 38.88 % of the complaints were documented on psilocybin, 33.33 % on MDMA, 18.51 % on LSD, and 9.25 % on ayahuasca. Up to 80% of the thirty-two adverse effects that occurred during a period spanning from week two up to twelve months included elevated blood pressure, headache, vomiting, feeling cold, panic attacks, transient anxiety, and depersonalization. Except for feeling cold, all of them had a large effect size (Fig. 2). According to the one-sample t-test, all four hallucinogens had a sizable effect size of reported adverse effects that was statistically significant (Fig. 3). The effect size based on the paired sample t-test yielded statistical significance for psilocybin (elevated blood pressure, headache, and panic attacks) (d = 3.739, 95% CI 1.697 to 5.763, p < 0.001) and MDMA (elevated BP, headache, panic attacks, and feeling cold) (d = 0.878, 95% CI 0.160 to 1.565, P= 0.016) but not for ayahuasca(headache) (d = 0.087, 95%CI -0.902-1.063, P = 0.873) or LSD (elevated BP and feeling cold) (d = 0.619, 95% CI-0.218 to 1.415, P = 0.153).

**Fig.2.**
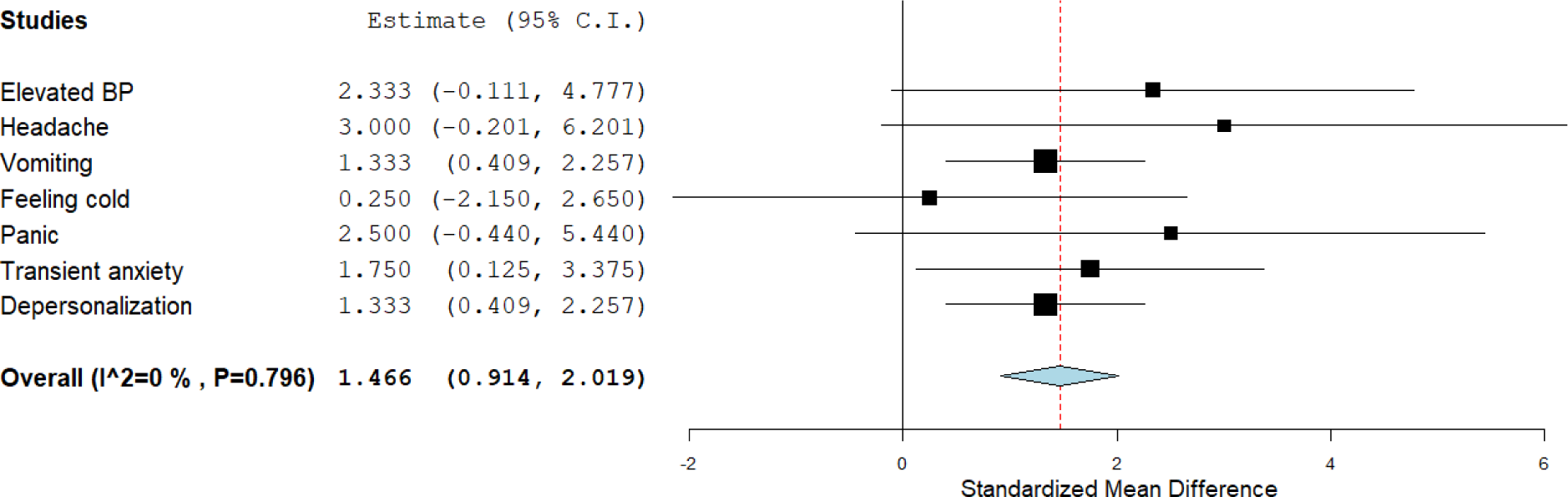
Up to 80% of the adverse effects and their effect size.

**Figure 3.**
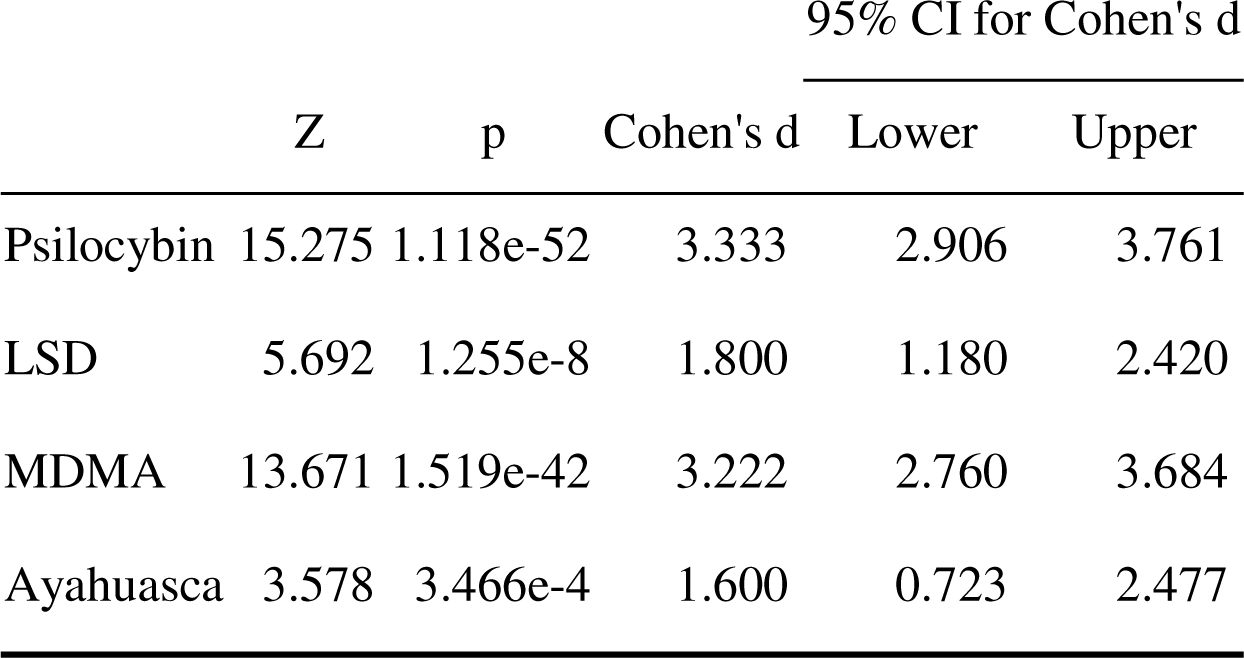
One Sample T-Test for the four hallucinogens.

## Discussion

### Summary of main results

Psilocybin may induce a rapid and sustained reduction in depressive and anxiety symptoms in severe depression and life-threatening cancer. Psilocybin may outperform citalopram at reducing depressive symptoms. A possible decrease in depressive symptoms was noted with MDMA, primarily in the long term in life-threatening cancer, autism spectrum disorder in adults, or PTSD. For anxiety symptoms, the effect size was either negligible or negative, with no statistical significance for MDMA. MDMA may reduce social anxiety symptoms. Ayahuasca may be effective at reducing depressive symptoms and LSD for anxiety in individuals with life-threatening cancer. The results of this review agree with those of Li and colleagues (38) and Goldberg and colleagues (41), indicating the benefits of psilocybin in reducing depressive symptoms in contrast to Galvo-Coelho and colleagues (43) who found no statistical significance of a benefit of psilocybin on depressive symptoms.

There were few reports of adverse effects in the selected trials, and data about dropout rates were scant. The paired sample t-test yielded statistical significance for psilocybin regarding elevated blood pressure, headaches, and panic attacks, and for MDMA regarding elevated blood pressure, headaches, panic attacks, and feeling cold.

### Overall completeness and applicability of evidence

Serotonin is linked to psilocybin, ayahuasca, LSD, and MDMA for managing depressive or anxiety symptoms. Hallucinogens influence 5-HT 1A, 5-HT 1C, 5-HT 2, and 5-HT3 to generate anxiolytic effects; 5-HT 1A and 5-HT2 generate antidepressant effects, and 5-HT3 generates reward and cognition improvement effects (65). In contrast, serotonin research provides no definitive answer to a relationship between serotonin and major depressive disorder or between low serotonin and anxiety or depressive symptoms (66). Hallucinogen’s pharmacological properties may be beyond serotonin. Indeed, psilocybin releases GABA and glutamate; LSD binds to D1 and D3 receptors; ayahuasca increases monoamine oxidase inhibitory (MAOI) characteristics; and MDMA releases oxytocin, vasopressin, and cortisol. Dopamine, NMDA, and rapamycin complex 1 (mTORC1) mediate ketamine’s therapeutic effects (15, 67). Therefore, the basic pharmacology of psilocybin, ayahuasca, LSD, and MDMA for anxiety and depression merits additional study. This brings together the importance of the validity of the depression-anxiety-serotonin paradigm.

The default mode network (DMN) connects brain areas associated with introspective activities, thinking, and autobiographical memory (68, 69). Posner and colleagues suggested that people with a high familial risk for major depressive disorder had more DMN connectivity and less negative DMN-central executive network (CEN) connectivity than individuals with a low familial risk (70). Schultz and colleagues suggested that decreased connectivity between the frontoparietal cognitive control network (FPN) and the rest of the brain is associated with depressive symptoms in the general population (71). Psilocybin reduces DMN connectivity (72), acute LSD administration reduces functional connectivity in the visual, sensorimotor, and auditory networks, and DMN connectivity (73), and ayahuasca lessens DMN brain activity (62). MDMA interferes with fear memory, resulting in various affiliative and prosocial behaviors (74). Psilocybin, ayahuasca, LSD, and MDMA may differ from commonly used medications to treat major depressive and anxiety disorders by influencing the default mode network (DMN) and suppressing fear and negative emotions, which could explain the possible large effect size.

Johnson and colleagues emphasized in a summary of hallucinogen studies and preventive measures that hallucinogens are not known to cause organ damage or neuropsychological deficiencies and that their toxicity is low. Despite being portrayed as physically safe and not a substance of dependence, hallucinogens can have psychological consequences like prolonged psychoses (75).

Dropout in RCTs is frequent and poses a risk to the validity of the data because completers and non-completers may have distinct characteristics (76). There is a great likelihood that individuals in experimental research will drop out if they experience unpleasant adverse effects. Psychological and physiological adverse effects were observed for psilocybin and MDMA. Clinical case reports on ayahuasca and LSD revealed troubling psychological effects. About 1–1.5 hours after ingesting ayahuasca, Rocha and colleagues (77) reported temporary disorientation (20–30 minutes), fear, anxiety, dissociation, depersonalization, agitation, confusion, anxiety, and visual hallucinations. Goldman and his colleagues wrote about a case of LSD flashback syndrome treated with an SSRI twenty-five years after the individual stopped using LSD (78). Despite the lack of significant adverse effects typically recorded, there is still cause for concern regarding the safety and tolerance of hallucinogens. Because of insufficient documentation on attrition, there is room to continue interrogating hallucinogens’ tolerability and safety.

### Abuse and misuse concerns

Psilocybin-containing mushrooms with varying concentrations are widely used illegally. Unprepared and unsupervised users may engage in harmful behavior, and individuals who already have a mental illness or are at risk of developing psychotic disorders may see their condition worsen (75). MDMA has the potential to cause physical dependency. Ecstasy use reflects “compulsive usage” and “escalating use” (37). LSD may accumulate in the body over time, resulting in user tolerance. LSD dependency can cause prolonged psychosis (79).

Using the Addiction Severity Index (ASI) (ASI Alcohol Use and Psychiatric Status subscales) on currently active ayahuasca users, Fábregas and colleagues found that ritual ayahuasca use did not seem to be linked to the significant psychosocial effects that are often caused by other drugs of abuse (80). Therefore, research on its potential for addiction merits further investigation.

### Limitations

Pre-selected standard measures were selected to gauge outcomes. Nevertheless, despite the extensive use of standardized scales in research, they may have limited therapeutic utility. Self-rating yielded more information than clinician-rating measures, which restricts the generalizability of this review’s findings.

While addressing the long-term effects of hallucinogens on depressive and anxiety symptoms, there was no control group. The absence of control groups can either inflate or deflate results. Hallucinogens were mainly used to facilitate psychotherapy adjuncts. The use of hallucinogens as psychotherapy adjuncts in most studies limits the interpretation of their actual benefits. Most trials enrolled a limited number of participants, and several were more experimental than clinical.

“Not all patients being compared have the same condition. Furthermore, some of the compounds included in this review have very different mechanisms of action. Drawing valuable conclusions from combining different conditions seems complicated. Clustering the analysis by a specific disease could have helped reduce confounders and generate a more accurate effect size. We use the term “hallucinogens” instead of “psychedelics” because not all the compounds reviewed fulfill the criteria of psychedelics. MDMA acts as both a stimulant and a hallucinogen (83).

Therefore, its effects may be more related to increased tenderness and contentment rather than changing perceptions. Most of the rating scale results were based on secondary analysis, which can significantly flaw the interpretation of the analysis.

## Conclusion

### Implications for practice and policy

Overall, psilocybin, MDMA, ayahuasca, and LSD may have the potential to assist in managing depressive and anxiety symptoms. Here in this review, the effect size appears to be significant. Despite a large effect size, arguments about hallucinogens’ potential therapeutic impact, risks for abuse and misuse, and the likelihood of psychological and physiological adverse effects need rigorous studies. The data we review are insufficient to support the use of psilocybin, MDMA, ayahuasca, or LSD to embed them in clinical practice. Further studies with broader sample sizes, clinically oriented, and the use of standardized scales that are clinically relevant are warranted.

### Implications for research

Hallucinogens affect vital brain areas related to major depressive and anxiety disorders. It seems that their pharmacological roots are beyond serotonin, this warrants further investigations. Much needs to be known about the long-term effects of psilocybin, ayahuasca, LSD, and MDMA on individuals with severe depression and anxiety disorders. Hallucinogens’ psychological consequences, such as prolonged psychoses, compared to their potential benefits for treating depressive and anxiety symptoms warrant investigations. Characteristics of completers and non-completers in the context of dropout in randomized controlled trials (RCTs) involving hallucinogens and how they might affect the validity of the results warrant further investigations.

## Data availability

The data availability criterion does not apply to this article because no new data were generated or analyzed for this investigation.

## Author contributions

The study was conceived and designed by D. F. D.F., V.K., and N.R. evaluated bibliographies, extracted data, analyzed the methodological quality of clinical trials, and then assigned JADAD scores. D.F. conducted the meta-analysis and drafted the initial version of the manuscript. D.F., V.K., and N.R. contributed to the writing of the manuscript and approved its final form.

## Funding

The author(s) received no financial support for the research, authorship, and/or publication of this article.

## Declaration of interest

The authors declare that they have no known competing financial interests or personal relationships that could have appeared to influence the work reported in this paper.

## Notes

### Competing Interest Statement

The authors have declared no competing interest.

### Clinical Protocols

https://www.crd.york.ac.uk/prospero/#myprospero

### Funding Statement

This study did not receive any funding

